# In-hospital Death Following Successful OHCA Resuscitation: Causes of early and late mortality and the impact of withdrawal of care

**DOI:** 10.1101/2020.03.15.20029207

**Authors:** Shu Li, Christos Lazaridis, Fernando D. Goldenberg, Atman P. Shah, Katie Tataris, David G. Beiser, Willard W. Sharp

## Abstract

**Objective:** In-hospital mortality in patients successfully resuscitated following out-of-hospital cardiac arrest (OHCA) is high. The factors and timings of these deaths is not well known. To better understand in hospital post-OHCA mortality we developed a novel categorization system of in hospital death and studied the factors and timings associated with these deaths.

**Methods:** This was a single-centered retrospective observational human study in adult non-traumatic OHCA patients in a university affiliated hospital. Through an expert consensus process, a novel classification system of hospital death was developed.

**Results:** Two hundred and forty-one patients were enrolled in the study. Death was categorized as due to withdrawal of life sustaining treatment (WOLST) 159 (66.0%), recurrent in-hospital cardiac arrest 51 (21.1%), or due to neurological criteria 31 (12.9%). Subcategorization of factors associated with WOLST into 7 categories was done by defined criteria. Inter-reliability of this system was 0.858. 50% of WOLST decisions were due to neurological injury. Early death (≤ 3 days) was associated with recurrent in-hospital cardiac arrest and WOLST in the setting of refractory shock or multi-organ injury. Late in-hospital death (> 3 days) was primarily due to WOLST decisions in the setting of isolated neurological injury.

**Conclusions:** OHCA in hospital mortality occurred in a bimodal pattern with early deaths due to recurrent arrest and multiorgan injury while late deaths were due to isolated neurological injury. The majority of deaths occurred in the setting of WOLST decisions. Further study of the influence of these factors on post OHCA survival are needed.

## INTRODUCTION

Each year, an estimated 320,000 adults experience Emergency Medical Services (EMS) assessed out-of-hospital cardiac arrests (OHCA) in the United States. Approximately 30% of patients resuscitated following OHCA are admitted to hospital, but less than 1/3 of these patients survive to hospital discharge (1). This high morbidity and mortality have been attributed to the post-CA syndrome associated with shock, inflammation, multi-organ injury, and neurological sequelae (2). Though targeted temperature management has improved post-CA outcomes, the pathophysiology of post-CA syndrome is not yet fully understood and additional therapies are needed.

To develop better therapies and systems for post-CA care, it is necessary to not only understand the factors associated with post-CA survival, but also those with non-survival. To date, there have been few studies characterizing causes of post-OHCA in-hospital death. Recent investigations demonstrate that many inpatient post-OHCA deaths are due to neurological injury and the withdrawal of life sustaining treatments (WOLST) (3-7). However, these studies have lacked a standardized approach to the categorization of injuries associated with post-OHCA deaths. In addition, the inter-rater reliability in these studies has been moderate and remains to be further improved. Here we present a novel classification system for post-OHCA in-hospital death with high inter-rater reliability that we used it to determine the timing and factors associated with in-hospital death in post-OHCA patients.

## MATERIALS AND METHODS

### Patients

This was a single-center, retrospective, observational cohort study between January 1, 2016 and August 31, 2019 at an urban tertiary medical center. OHCA victims who had successful return of spontaneous circulation (ROSC) in the emergency department were identified, based on following criteria: (1) Age ≥ 18 years old; (2) Non-traumatic out-of-hospital cardiac arrest; (3) Achieved sustained ROSC (>20 minutes); (4) Death prior to hospital discharge. The study protocol received a waiver of informed consent from the Institutional Review Board.

Targeted temperature management was started immediately following ROSC either in ED or ICU according to hospital-wide protocol and following American Heart Association guidelines (2). Patients surviving to admission received routine intensive care unit (ICU) care. The declaration of death by neurological criteria was made by a neuro-intensivist, following criteria from the 2010 American Academy of Neurology guideline (8). Data was collected from electronic medical records including demographics of the patients and characteristics of the initial cardiac arrest events as well as post-arrest treatment.

### Definition of Categories

In consultation with an expert panel of specialists in Emergency Medicine, Neurology Intensive Care and Cardiology, a categorization system of post-CA death was developed (Supplementary Table 1). Three major categories of death were identified including recurrent in-hospital cardiac arrest (R-IHCA), death by neurological criteria (D-NC) or withdrawal of life-sustaining treatment (WOLST). In cases of WOLST, patients were further classified into one of seven categories dependent upon their medical conditions at time of WOLST. These categories included Refractory shock, Refractory respiratory failure/hypoxia, Neurological injury, Multi-organ injury, Pre-morbid conditions, Unknown prior *Do-Not-Resuscitate* (DNR) status and Indeterminate. The median time to death was set as the dividing point for Early and Late death.

### Inter-rater Reliability of Categorization

To assess inter-rater reliability, ten medical records were reviewed independently by two physicians (SL and WWS) who assigned each case to one of the death categories and a Kappa score was then calculated. Further review of the category definitions was then performed and the methodology used to acquire patient information was refined. This process was then repeated with another two sets of ten sequential cases with the intent of refining the category definitions until they were clear and substantial agreement was achieved. Once this had been done, the remaining patients in the cohort were then categorized and a final Kappa score was determined. (Figure 1). In cases of disagreement, the case was to be re-reviewed by the two reviewers who would discuss the discrepancy and determined the final classification of the cases. In cases of possible disagreement between the two reviewers, a third reviewer was available to review the case.

**Figure 1:**
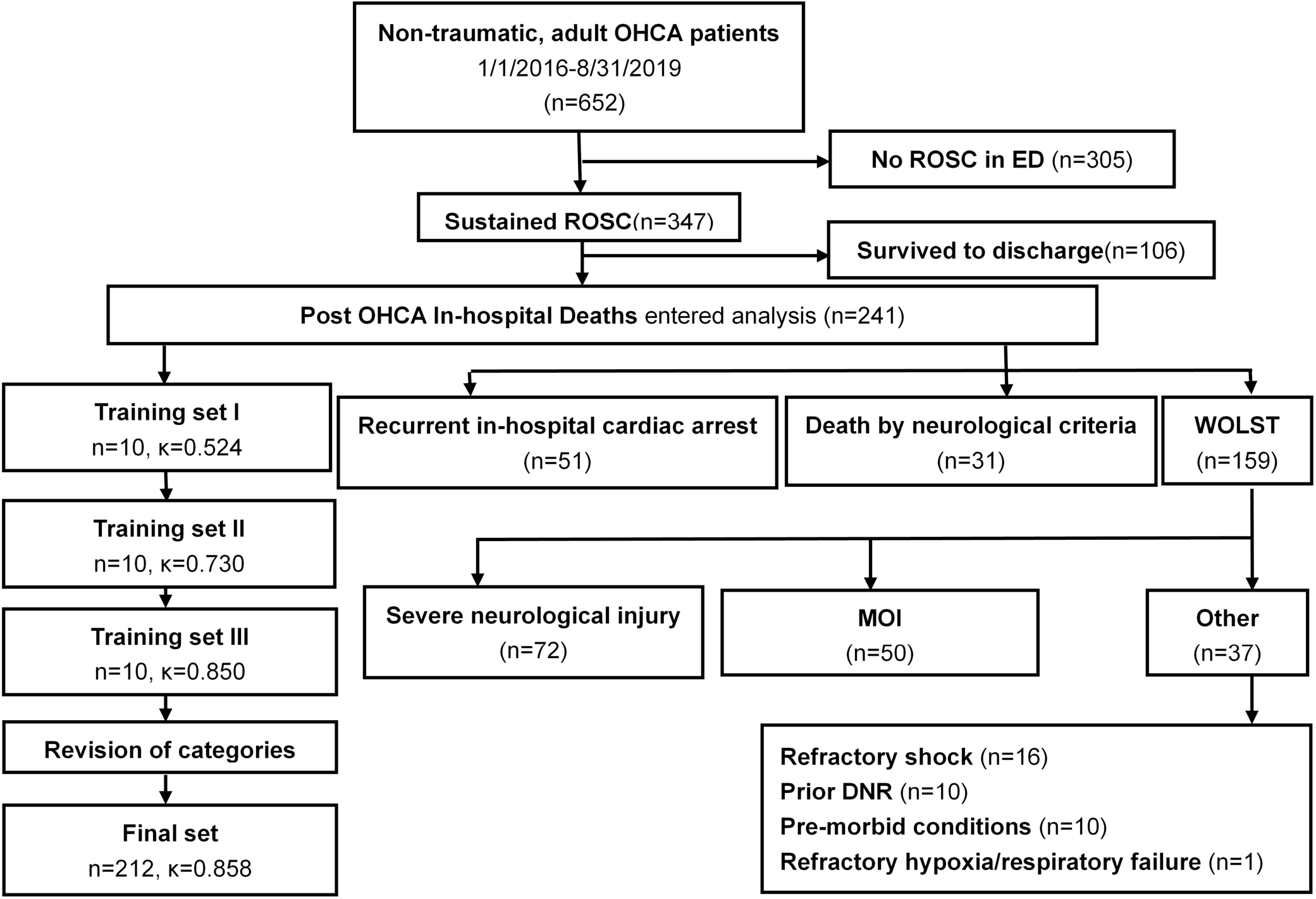
Patient population, categories and Kappa for inter-rater reliability.

### Statistical Analysis

The inter-rater reliability was assessed using Fleiss’ Kappa. A Kappa below 0.60 was considered moderate agreement, 0.61-0.80 was considered substantial agreement and Kappa above 0.81 was considered near perfect. Median with interquartile range (IQR) was used for continuous variables and counts and frequencies for categorical variables. Continuous data were compared with Mann-Whitney Test, and categorical data with Chi-Square Test or Fisher’s exact test. Variables were identified in univariate analyses with a *p* value of less than 0.05 or those that were considered of clinical significance, including age, gender, race, history of cancer, history of substance abuse, duration of CPR, pre-hospital ROSC, initial lactate level, and post-ROSC vasopressor dependence. Multivariable analyses were then performed using Binary logistic regression. A multivariable formula was generated through backward stepwise selection by Maximum Likelihood Estimation. All analyses were two-sided with a significance level of 0.05 and performed using IBM SPSS Statistics software, version 22.0 for Windows.

## RESULTS

From January 2016 to August 2019, there were 652 adult non-traumatic cardiac arrests transported to the emergency department. Of these 347 (53.2%) patients achieved sustained ROSC (> 20 minutes). There were 106 patients (30.5%) survived to hospital discharge with 47(13.5%) having a favorable neurological outcome (cerebral performance category score at 1-2). Our study then used the 241 (69.5%) of patients who died in the hospital for analysis (Figure 1).

The median age was 64 (IQR 54,78) years. Approximately half (49.4%) of the cases occurred in males. 85.1% of the patients were African-American and 36(15.7%) patients were reported to have history of substance abuse. Witnessed arrests occurred in 166 (68.9%) patients, but only 61 (25.3%) patients received bystander CPR. Cardiac arrest cause was presumed to be due to cardiac origin by EMS in 93 (58.5%) cases while 30 (12.4%) of the arrests presented with shockable rhythm. Other patient characteristics are listed in Table 1.

**Table 1.**
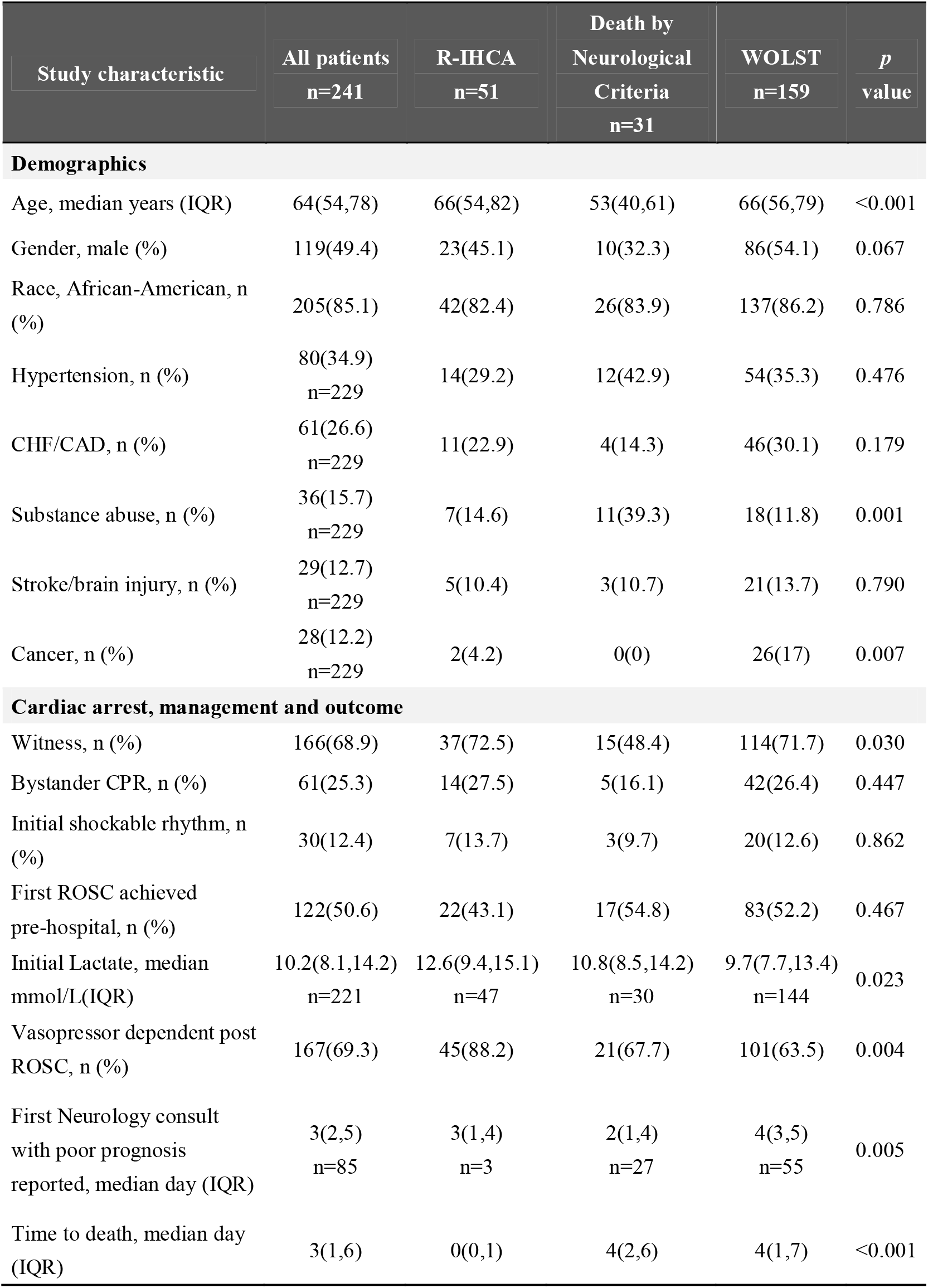
Patient demographics, arrest characteristics and post arrest treatment for different cause of death (Different cause of death)

### Inter-rater reliability

After initial review of the first ten cases in training set I, the Kappa between the two reviewers was 0.524. Discussion and refinement of the category definitions was performed, and the process was repeated using another ten patients for training sets II and III with Kappa of 0.730 and 0.850 respectively. Final category definitions were then used to categorize the remaining 211 patients with a final Kappa of 0.858 (Figure 1). Discrepancy between the two reviewers was 11.4% (Supplemental Table 2). Consensus was reached without the need for introducing the third reviewer.

### Cause of death

Post OHCA in-hospital deaths were classified as withdrawal of life sustaining treatments (WOLST) 159 (65.9%), recurrent in hospital cardiac arrest (R-IHCA) 51(21.2%), and death by neurological criteria (D-NC) 31(12.9%). Age, comorbidities, witnessed arrest, initial lactate level, post-ROSC vasopressor dependence and time of unfavorable neurological prognosis were found to be different among the three groups (Table 1).

R-IHCA patients differed from D-NC and WOLST patients by earlier DNR orders (0 vs. 3.5 and 2 days, p = 0.001 and p < 0.001, respectively).They also had significantly higher initial lactate levels (12.6 vs. 9.7 mmol/L, p = 0.008), and higher rates of vasopressor dependence post-ROSC (88.2% vs. 63.5%, p = 0.001) than WOLST patients. When compared to R-IHCA and WOLST patients, D-NC patients tended to be younger (53 vs. 66 and 66 years, p < 0.001 and < 0.001, respectively), had higher rates of substance abuse history (39.3% vs. 14.6% and 11.8%, p = 0.015 and 0.001, respectively), and higher rates of acute ischemic findings on brain computer tomography (CT) scans (82.8% vs. 40% and 34.4%, p = 0.003 and p < 0.001, respectively) while sharing similar comorbid burdens. D-NC patients were also less likely to have been witnessed arrests (48.4% vs. 71.7%, p = 0.001) and had earlier unfavorable neurological consult prognosis (2 vs. 4 days, p = 0.001) when compared to patients who had WOLST.

WOLST decisions were made primarily in the context of Neurologic injury 72 (45.3%), Multi-organ injury 50 (31.4%), Refractory shock 16 (10.1%), Prior DNR order 10 (6.3%) or Pre-Morbid conditions 10 (6.3%). Age, initial lactate, rate of initial shockable rhythm, and post-ROSC vasopressor dependence were also found to be different among subgroups (Table 2).

**Table 2.**
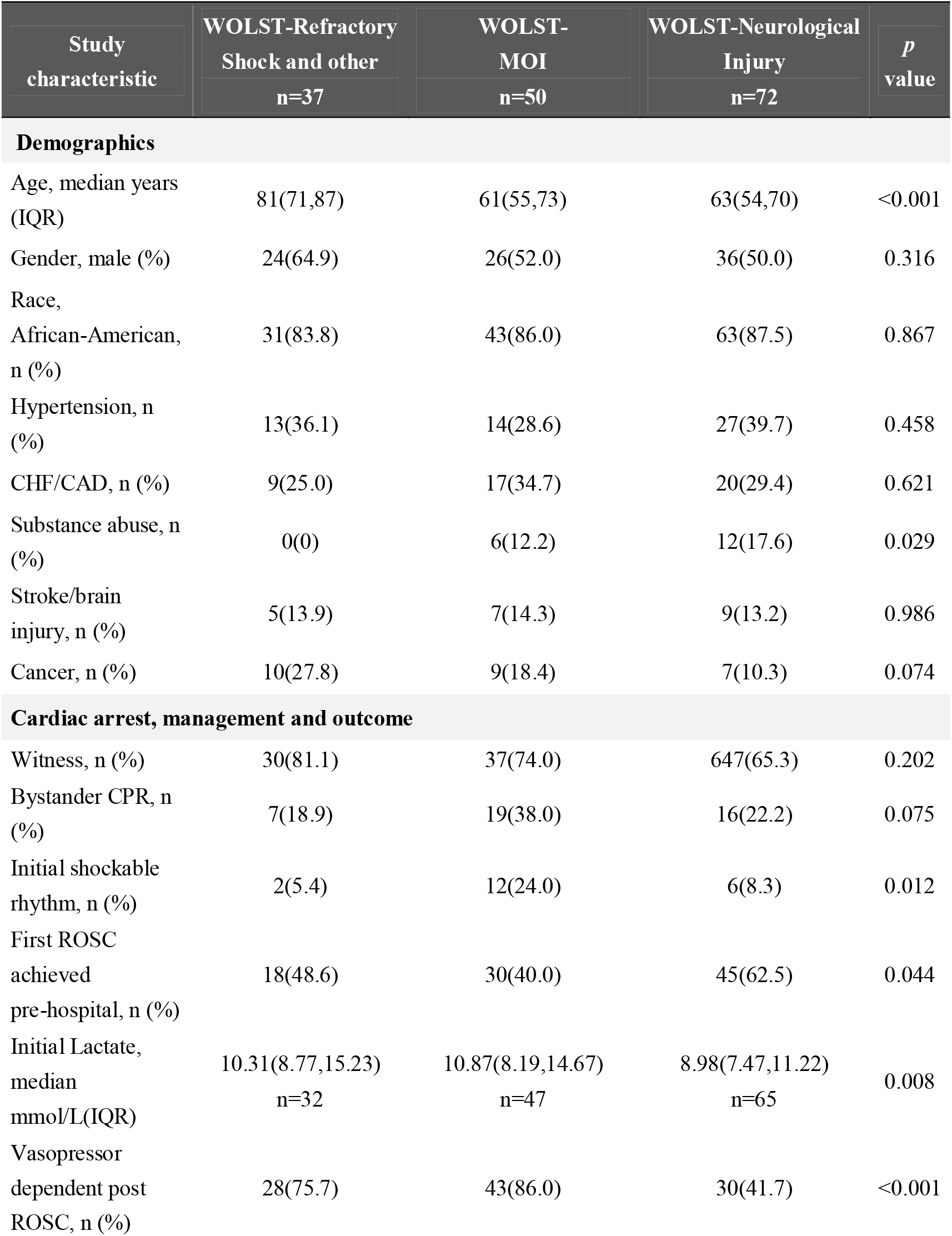
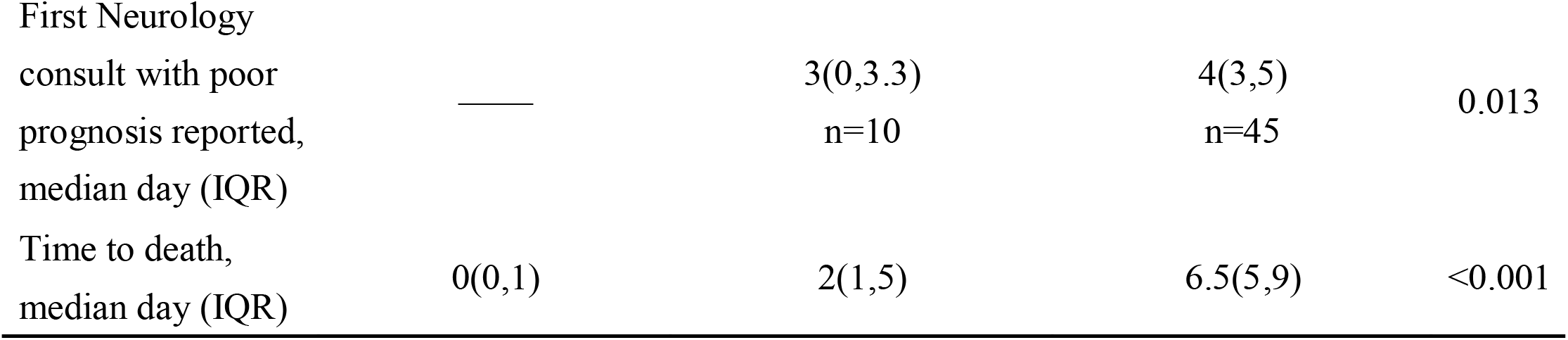
Demographics, arrest characteristics and post arrest treatment for patients of different conditions at time of withdrawal of life-sustaining treatment (Different Conditions at time of withdrawal of life-sustaining treatment)

### Time of death

The median time to death for the entire patient population was hospital day three. This median time to death was set as the dividing point for categorization of death as Early or Late death. R-IHCA patients had shorter median time of death (day 0, IQR 0,1) compared to WOLST patients (day 4, IQR 1,7) and D-NC patients (day 4, IQR 2,6) (Figure 2A). The daily number of deaths declined as the length of hospitalization increased (Figure 2B).

**Figure 2:**
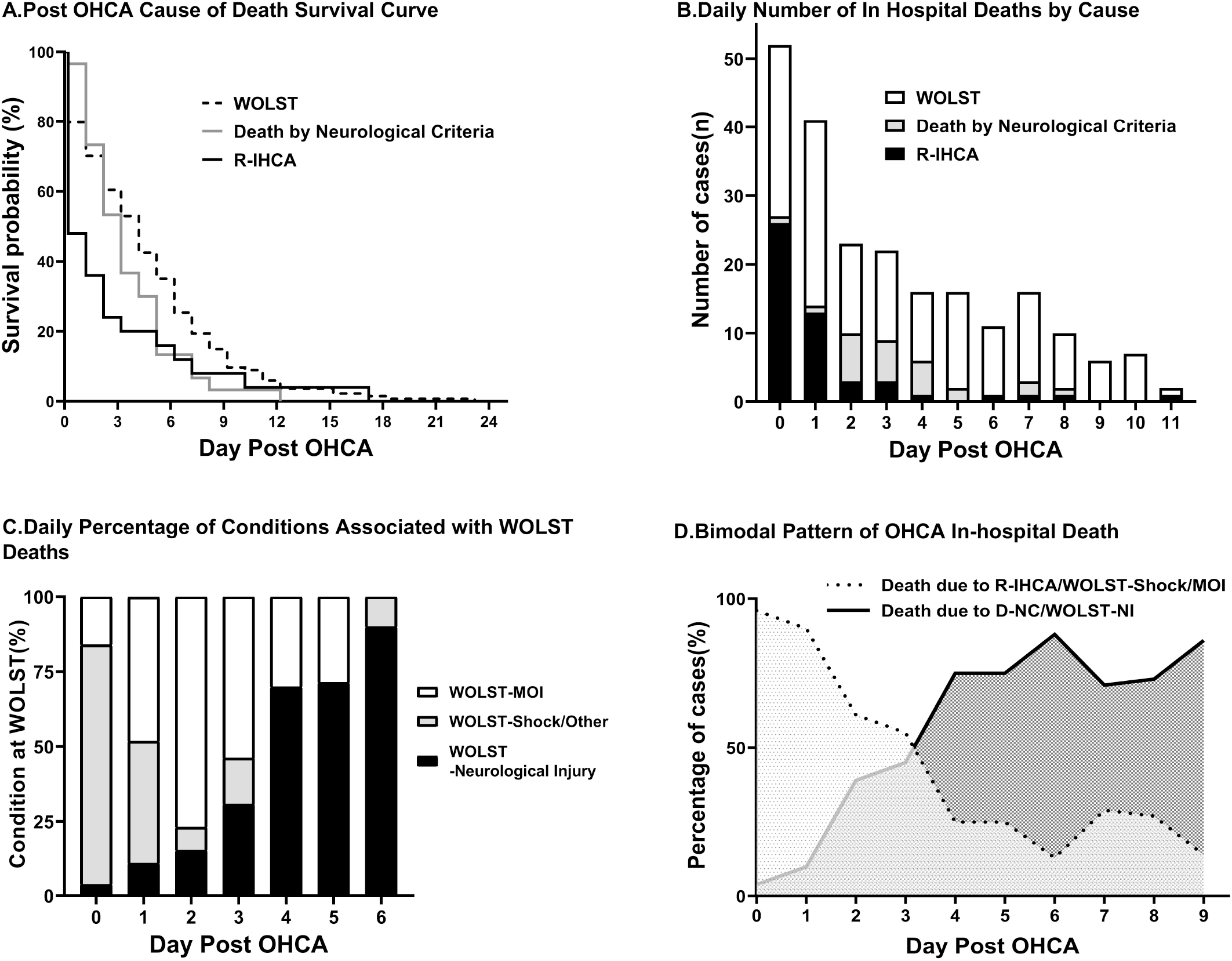
Death of post cardiac arrest patients with return of spontaneous circulation. **(A)** Kaplan Meier survival curves of different cause of death **(B)** Daily number of in hospital deaths by cause. **(C)** Daily percentage of in hospital deaths by cause **(D)** Bimodal pattern of Post OHCA in hospital death

Deaths prior to day three (Early death) were primarily categorized as R-IHCA 45 (32.6%), Multi-organ injury (WOLST-MOI) 34 (24.6%) or WOLST due to Refractory shock (WOLST-RS) 14 (10.1%). However, the percentage and number of deaths in these categories declined thereafter while in contrast daily hospital death in the categories of WOLST with Neurological injury (WOLST-NI) 62 (60.2%), Death by neurological criteria 16 (15.5%) increased. As expected, age, history of substance abuse, bystander CPR, initial lactate level, rate of post-ROSC vasopressor dependence and unfavorable neurological prognosis were also found to be different between Early and Late deaths (Table 3, Figure 2C).

**Table 3.** Demographics, arrest characteristics and post arrest treatment for patients with Early and Late death (Early and Late death)

| Study characteristic | All patients<br>n=241 | Early death<br>n=138 | Late Death<br>n=103 | *p*<br>value |
| --- | --- | --- | --- | --- |
| **Demographics** |  |  |  |  |
| Age, median years (IQR) | 64(54,78) | 67(56,81) | 62(51,70) | 0.002 |
| Gender, male (%) | 119(49.4) | 70(50.7) | 49(47.6) | 0.628 |
| Race, African-American, n (%) | 205(85.1) | 116(84.1) | 89(86.4) | 0.613 |
| Hypertension, n (%) | 80(34.9)<br>n=229 | 44(33.1) | 36(37.5) | 0.489 |
| CHF/CAD, n (%) | 61(26.6)<br>n=229 | 34(25.6) | 27(28.1) | 0.665 |
| Substance abuse, n (%) | 36(15.7)<br>n=229 | 14(10.5) | 22(22.9) | 0.011 |
| Stroke/brain injury, n (%) | 29(12.7)<br>n=229 | 15(11.3) | 14(14.6) | 0.458 |
| Cancer, n (%) | 28(12.2)<br>n=229 | 20(15) | 8(8.3) | 0.126 |
| **Cardiac arrest, management and outcome** |  |  |  |  |
| Witness, n (%) | 166(68.9) | 102(73.9) | 64(62.1) | 0.051 |
| Bystander CPR, n (%) | 61(25.3) | 42(30.4) | 19(18.4) | 0.034 |
| Initial shockable rhythm, n (%) | 30(12.4) | 20(14.5) | 10(9.7) | 0.266 |
| First ROSC achieved pre-hospital, n (%) | 122(50.6) | 80(58.0) | 39(37.9) | 0.002 |
| Initial Lactate, median mmol/L(IQR) | 10.2(8.1,14.2)<br>n=221 | 11.7(8.6,14.6) | 9.1(7.6,12.2) | <0.001 |
| Vasopressor dependent post ROSC, n (%) | 167(69.3) | 113(81.9) | 54(52.4) | <0.001 |
| First Neurology consult with poor prognosis reported, median day (IQR) | 3(2,5)<br>n=85 | 2(1,3) | 4(3,5) | <0.001 |
| Time to death, median day (IQR) | 3(1,6) | 1(0,2) | 7(5,9) | <0.001 |

Multivariable logistic regression model was built with age (OR =1.032, 95% CI =1.011–1.053), initial lactate level(OR =1.122, 95% CI =1.027–1.225), duration of CPR exceeding 30 minutes(OR =2.915, 95% CI =1.467–5.791), and vasopressor dependence post ROSC(OR =3.872, 95% CI =1.904–7.875) as the dependent variables for Early death during hospital stay.

## DISCUSSION

### Post-OHCA In-Hospital Death Categorization

In this study, we developed a novel system of categorizing post-OHCA in-hospital death to determine the timing and factors associated with post-resuscitation mortality. Inter-rater reliability of this system was near perfect (0.858) with none of the cases being classified as indeterminate. Using this classification system, we determined that the majority of post-CA in-hospital deaths in our cohort occurred in the context of WOLST decisions and that nearly half of these decisions occurred in the context of isolated neurological injury. Our study is not the first to attempt to categorize the mode of in-hospital death following OHCA. Herlitz et al. originally attempted to categorize post OHCA deaths as due to brain damage (45%), myocardial damage (9%), or both (40%) (9). This prior study revealed that neurological injury was common in OHCA patients but was significantly limited in its patient population studying only patients presenting with ventricular fibrillation. It also lacked well-defined death categories and neglected the impact of other organ injuries as well as WOLST decisions on survival. Lemiale et al. also attempted to characterize post-CA deaths in ICU setting, categorizing death as due to post-arrest shock 269 (34.8 %) or neurological injury 499 (65.2 %) (10). However, it also failed to address the probability of the co-existence of both injuries and did not address the impact of WOLST on outcomes. More recently, Witten et al. developed a system to systematically categorize post-OHCA in-hospital deaths using more defined criteria representing substantial progress. However, it did not distinguish between patients dying in the setting of WOLST decision despite the high prevalence of WOLST and its complex effects on patient outcomes (11). The inter-rater reliability rate of 0.61 was substantial. It was likely less than ideal due to a lack of strict objective criteria for some of the categories. In addition, the presence of a “miscellaneous category” for undefined patients, may have led to bias for the results of statistical analysis.

In contrast to these previous studies, our study had nine categories for classifying post-OHCA in-hospital death and was based on defined elements of organ injury. These categories were determined by requirements for objective evidence and exclusive criteria for each category rather than relying on subjective determinations of injury simply based on documentation in chart or judgement of reviewers. In addition, none of the cases were classified as Indeterminate and our study reached higher inter-rater reliability than previous studies.

### Cause of Death

In our study, WOLST decisions accounted for nearly two thirds of all deaths and predominated as the cause of death at any given day post-cardiac arrest. Our findings are similar to those of Albaeni and Witten demonstrating that 64% and 77%of deaths respectively occurred in the context of treatment withdrawal (3, 11). In contrast, re-current cardiac arrest was a significant factor for death on the initial day of hospitalization (Day 0) but by hospital day one (Day 1) had significantly declined thereafter. Death by neurological criteria was largest on hospital Day 2 and Day 3 and was only a small percentage thereafter.

It was impossible to determine why caregivers and families made decisions regarding WOLST, but these decisions were often guided by evidence of organ injury. On Day 0, WOLST decisions were made primarily in the context of Refractory shock as evidenced by the utilization of multiple vasopressors. Post-cardiac arrest hemodynamic shock was common and likely contribute not only to the high rate of re-arrest but also to the WOLST decisions during this period. However, WOLST decisions in the setting of shock dropped to 18.5% by day 1 and 7.7% by day 2 with an increasing number of WOLST decisions being made in the setting of multi-organ injury, which peaked by Day 2 and gradually decreased thereafter. Finally, WOLST decisions in the context of neurological injury with objective evidences and the lack of other major organ injuries increasingly predominated WOLST decisions on post OHCA Day 4 onwards.

### Timing of Death

In-hospital deaths were noted to follow a biphasic pattern centered on the median day of death (hospital day 3). Early deaths (<day 3) occurred primarily due to recurrent cardiac arrest or WOLST decisions in the context of hemodynamic shock, or multi-organ injury. In contrast late deaths (>day 3) occurred primarily due to WOLST decisions in the context of neurological injury (Figure 2D). These finding demonstrate several important things. First, patient care in compliance with post-arrest therapeutic hypothermia clinical guidelines is likely heavily influencing the median day for hospitalization. Current guidelines stipulate that neuro-prognostication should not occur until 72 hours following targeted temperature management (Post OHCA day 4) which is when a large percentage of WOLST decisions due to neurological injury begin to occur. Thus, any alteration in current guidelines or hypothermia duration will impact the timing of post-OHCA survival. Second, it is interesting to speculate to whether these two patient populations of early and late deaths represent two different injury patterns or whether they simply represent a continuum of post-cardiac arrest injury. Our findings would suggest both of these conclusions are true. Nearly 42% of patients who ultimately died due to WOLST decisions in the context of neurological injury had been on vasopressor support during the initial few days of their hospitalization while 58% of these patients had no vasopressor requirements at the time of their admission. Further study of these two patient populations is needed to determine if there are pathophysiological differences which could impact future patient treatment studies.

### Factors Associated with Death

To date, there have been some studies on risk factors of hospital mortality and long-term prognosis for OHCA patients which indicate that advanced age, comorbidities and increased lactate measurements were associated with a higher risk of in-hospital death for both shock and brain injury (10, 12, 13). However, few studies have evaluated the factors associated with different times and causes of post-OHCA in-hospital death. Thus, we also performed multivariate modeling of our data and confirmed that longer durations of CPR prior to sustained ROSC, post-ROSC vasopressor dependence, and initial lactate level were predictors of Early death.

### Post-CA outcomes in a primarily African American population

Previous studies of OHCA have been reflective of predominately Caucasian populations (64% to 81.2%) (3, 11, 13-15). In contrast, our study consisted predominately of African Americans (86.8%) making our study cohort unique. Rates of sudden cardiac arrest have been reported to be significantly higher in African-Americans (16). This has been attributed to the lack of access to health care, less bystander CPR (17) and lower overall rates of survival to hospital discharge compared with Caucasian patients (15, 18). In addition, there are socioeconomic and genetic basis for racial outcomes (17). Despite these pre-disposing factors, overall ROSC rates for OHCA patients transported to our hospital were 43.8% and overall survival was 18.4%, comparing favorably to the national ROSC and survival rates of 43.8% and 16.1% (19).

### Limitations

There are several important limitations of our study. First, this was a single-center, retrospective study enrolling only 241 patients. Further studies using our categorization system in a higher number of patients and other hospital settings will be necessary before broader conclusions can be drawn. Second, the timing and factors associated with post-CA death in our study could have been influenced by our patient population which consisted pre-dominantly of African American patients. This population has not been well studied for OHCA and is different from the racial backgrounds of other typical OHCA studies. Further study of post-CA outcomes on our patient population is needed.

## CONCLUSION

In this study we developed a novel nine-category system for categorizing in-hospital death following OHCA resuscitation that had high inter-rater reliability. Applying this system retrospectively to a cohort of OHCA patients, we determined that the majority of in-hospital deaths were associated with WOLST. Death in the first three days of hospitalization (Early) were most often due to recurrent cardiac arrest or WOLST associated with recurrent shock or multi-organ injury. In contrast, most deaths after 3 days (Late) were due to WOLST in the context of isolated neurological injury. We conclude that post-OHCA in hospital death is characterized by a bi-modal pattern of organ injury and death and that these outcomes may be heavily influenced by WOLST decisions.

## Data Availability

To facilitate this study and to monitor quality of care, a REDCAP HIPPA secure registry was maintained with EMR as source data.

## ACKNOWLEDGEMENTS

We are grateful for Dr. Michael Millis and Dr. Mark Siegler from the University of Chicago Medicine for their proposal of the fellowship and great support provided through the whole academic year. Special thanks to Dr. Jing Xie Ph.D. from Peking Union Medical College Hospital for all the critical advisement for statistical analysis.

